# Technical Development and In Silico Implementation of SyntheticMR in Head and Neck Adaptive Radiation Therapy: A Prospective R-IDEAL Stage 0/1 Technology Development Report

**DOI:** 10.1101/2024.08.29.24312591

**Authors:** Lucas McCullum, Samuel Mulder, Natalie West, Robert Aghoghovbia, Alaa Mohamed Shawky Ali, Hayden Scott, Travis C. Salzillo, Yao Ding, Alex Dresner, Ergys Subashi, Dan Ma, R. Jason Stafford, Ken-Pin Hwang, Clifton D. Fuller

**Affiliations:** UT MD Anderson Cancer Center UTHealth Houston Graduate School of Biomedical Sciences, Houston, USA; Department of Radiation Oncology, The University of Texas MD Anderson Cancer Center, Houston, TX, USA; Morehouse School of Medicine, Atlanta, USA; Department of Radiation Physics, The University of Texas MD Anderson Cancer Center, Houston, TX, USA; Philips Healthcare MR Oncology, Cleveland, Ohio, USA; Department of Biomedical Engineering, Case Western Reserve University, Cleveland, Ohio, USA; Department of Imaging Physics, The University of Texas MD Anderson Cancer Center, Houston, TX, USA

**Keywords:** SyntheticMR, SyMRI, MRI, Quantitative Imaging, MR-Linac, Radiotherapy, Radiation Therapy, Adaptive Radiation Therapy, Head and Neck Cancer

## Abstract

**Objective:** The purpose of this study was to investigate the technical feasibility of integrating the quantitative maps available from SyntheticMR into the head and neck adaptive radiation oncology workflow. While SyntheticMR has been investigated for diagnostic applications, no studies have investigated its feasibility and potential for MR-Simulation or MR-Linac workflow. Demonstrating the feasibility of using this technique will facilitate rapid quantitative biomarker extraction which can be leveraged to guide adaptive radiation therapy decision making.

**Approach:** Two phantoms, two healthy volunteers, and one patient were scanned using SyntheticMR on the MR-Simulation and MR-Linac devices with scan times between four to six minutes. Images in phantoms and volunteers were conducted in a test/retest protocol. The correlation between measured and reference quantitative T1, T2, and PD values were determined across clinical ranges in the phantom. Distortion was also studied. Contours of head and neck organs-at-risk (OAR) were drawn and applied to extract T1, T2, and PD. These values were plotted against each other, clusters were computed, and their separability significance was determined to evaluate SyntheticMR for differentiating tumor and normal tissue.

**Main Results:** The Lin’s Concordance Correlation Coefficient between the measured and phantom reference values was above 0.98 for both the MR-Sim and MR-Linac. No significant levels of distortion were measured. The mean bias between the measured and phantom reference values across repeated scans was below 4% for T1, 7% for T2, and 4% for PD for both the MR-Sim and MR-Linac. For T1 vs. T2 and T1 vs. PD, the GTV contour exhibited perfect purity against neighboring OARs while being 0.7 for T2 vs. PD. All cluster significance levels between the GTV and the nearest OAR, the tongue, using the SigClust method was p < 0.001.

**Significance:** The technical feasibility of SyntheticMR was confirmed. Application of this technique to the head and neck adaptive radiation therapy workflow can enrich the current quantitative biomarker landscape.

## 1. Introduction

Magnetic resonance imaging (MRI) plays a vital role in visualizing tissues which are often indistinguishably isointense on computed tomography (CT)^1^. This becomes especially important in the field of radiation oncology due to most malignancies being in the soft tissue where CT has lower contrast. Due to this superiority, systems designed for radiation therapy (i.e., the linear accelerator, or Linac) have integrated diagnostic-level MRIs to create the 1.5T MR-Linac^2^, allowing for simultaneous radiation therapy delivery and MRI acquisition for anatomically dynamic structures (i.e., the lung). Furthermore, due to the availability of imaging acquired at each treatment fraction, strategies to adapt the treatment delivery based on imaging changes have gained traction and is known as adaptive radiation therapy, or ART.

However, unlike CT, the radiation therapy workflow in MRI is currently focused around qualitative rather than quantitative representations of tumor and healthy tissue using the on-board MRI system of the MR-Linac. Though easy to interpret visually, the arbitrary signal values associated with standard relaxation “weighted” images in MRI present hurdles to the standard radiation therapy workflow, not the least of which are that these images are not ideal to monitor treatment response since they do not provide consistent quantitative measurements which can be compared across the different fractions of treatment^3^. In head and neck cancer, quantitative MRI probing tissue T1 and T2 properties has recently been investigated for diagnosis^4^, assessment of treatment response^5^, and assessment of normal tissue damage^6^. However, on the 1.5T MR-Linac, T1 and T2 mapping has been limited to acquisition times of exceeding 3 minutes and 5 minutes, respectively^7,8^. Therefore, acquiring T1 and T2 maps alone will require upwards of 8 minutes and suffer from higher misregistration error due to the potential for patient adjustments intra- and inter-scan. This time requirement is critical on the MR-Linac where, typically, less than 10 minutes are available for elective imaging within the clinical workflow^9^, and this will only decrease as broader patient coverage is demanded with the recently introduced Comprehensive Motion Management (CMM) software^10^.

The company SyntheticMR (Linköping, Sweden) has developed a single scan time (i.e., <6 minutes) simultaneous multiparametric MRI acquisition sequence originally known as QRAPMASTER^11^ and more regularly known as multi-dynamic multi-echo (MDME) on Siemens MRI scanners, MAGiC on GE scanners, and SyntAc on Philips scanner. Through their post-processing software, SyMRI, these acquired images can be reconstructed to quantitative T1, T2, and proton density (PD) maps and derivative synthetic contrast maps such as T1/T2/PD-weighted and inversion recovery (IR), e.g., fluid attenuated (FLAIR), phase-sensitive (PSIR), and short inversion time (STIR). The most common implementation of the sequence is the 2D-MDME^11–13^ which acquires multiple 2D slices, often with slice thicknesses of 3 – 6 mm while in-plane resolution is often <2 mm. However, a 3D version of the sequence capable of 1 mm isotropic acquisitions (3D-QALAS)^14^, has been developed and has shown clinically acceptable quantitative accuracy and repeatability in a multi-center^15^ and multi-vendor study^16^. More details of the MR physics, technical considerations, and pulse sequence design can be seen in the review article by Hwang et al. 2022^17^.

SyntheticMR has seen increasing usage for diagnostic imaging^18–21^, however limited investigation has been conducted in usage for radiation oncology^22–25^ and even fewer studies have focused on the head and neck^26,27^. Further, no studies to the author’s knowledge have evaluated the technical feasibility of the SyntheticMR sequence on MR-Simulation (MR-Sim) or MR-Linac devices. The potential application of SyntheticMR to the radiation oncology space has high promise for increasing the dimensionality available for treatment monitoring and optimal adaptive therapy decisions. Specifically, within the head and neck oncology space, due to the large number of organs-at-risk (OARs) near the target, advanced quantitative MRI techniques would be advantageous to characterize simultaneous normal tissue dose-responses and tumor control more effectively. Therefore, the purpose of this study is to investigate the technical feasibility of integrating SyntheticMR into the head and neck adaptive radiation therapy workflow on both an MR-Sim and MR-Linac scanner. This will be presented using the radiotherapy-predicate studies, idea, development, exploration, assessment, and long-term study (R-IDEAL) framework, as recommended by the MR-Linac Consortium, completing Stage 0 (radiotherapy predicate studies) and Stage 1 (first time use) systematic evaluations^28^.

## 2. Methods and Materials

### 2.1. MRI Acquisition Parameters

To evaluate SyntheticMR across the radiation oncology department, MRI scans were performed on a 3T Siemens Vida MR-Sim scanner (Siemens Healthcare; Erlangen, Germany) and a 1.5T MR-Linac (Unity; Elekta AB; Stockholm, Sweden). For this study, the 2D-MDME sequence was acquired with acquisition parameters as shown in Table 1. These parameters were chosen to optimize the acquisition for different clinical scenarios on the MR-Linac including the possibility for lymph node evaluation^29^ (coarse sequence) and required resolution for stereotactic radiation therapy precision^30^ (fine sequence). Further, due to the non-isotropic acquisition of 2D-MDME, all scans were acquired in the axial / transverse orientation to best visualize structures in the head and neck at all points in the radiation therapy workflow^31,32^.

**Table 1.** Acquisition parameters across all scanners utilized in this study.

|  | 3T Siemens Vida | 1.5T Elekta Unity<br>(coarse sequence) | 1.5T Elekta Unity<br>(fine sequence) |
| --- | --- | --- | --- |
| **Software Version** | XA50 | R5.7.1.2 | R5.7.1.2 |
| **Field-of-View (mm<sup>3</sup>)** | 256 x 256 x 149 | 250 x 349 x 199 | 256 x 256 x 179 |
| **Acq. Voxel Size (mm<sup>3</sup>)** | 1.00 x 1.67 | 2.02 x 2.65 | 1.00 x 1.03 |
| **Recon. Voxel Size (mm<sup>3</sup>)** | 0.5 x 0.5 | 0.99 x 0.99 | 0.5 x 0.5 |
| **Slice Thickness (mm)** | 4 | 3 | 4 |
| **Slice Gap (mm)** | 1 | 1 | 1 |
| **Number of Slices** | 30 | 50 | 36 |
| **TR / TE<sub>1</sub> / TE<sub>2</sub> (ms)** | 4330 / 19 / 94 | 8599 / 11 / 102 | 6191 / 12 / 112 |
| **Echo Train Length (ETL)** | 12 | 12 | 12 |
| **Acceleration**<br>**(GRAPPA / CS-SENSE)** | 3 | 3 | 3 |
| **Refocusing Flip Angle (°)** | 150 | 180 | 180 |
| **Acquisition Time (m:ss)** | 4:06 | 4:44 | 5:53 |

### 2.2. MR Phantoms Assessed

The American College of Radiology (ACR) large phantom was used to evaluate geometric distortion and accuracy. The CaliberMRI “ISMRM/NIST” Premium System Phantom Model 130 phantom (CaliberMRI; Boulder, CO) was used as a reference for NIST-traceable T1, T2, and PD values^33^. This phantom includes 14 vials for each metric (42 total vials) suitable for T1 values between 20 and 1724 ms, T2 values between 9 and 853 ms, and PD values between 5 and 100% at 20°C on a 1.5T MRI scanner. This was confirmed during routine quality assurance that the temperature inside the bore averaged 20°C with minimal fluctuations. Reference values are also provided by the CaliberMRI for 3T MRI scanners. To evaluate repeatability, two repetitions in phantom were performed in a test-retest method using a coronal slice orientation. Similarly, to evaluate reproducibility, this process was reproduced across both the MR-Sim and MR-Linac scanners.

### 2.3. Healthy Volunteer and Patient Description

Two healthy volunteers were imaged on both the MR-Sim and MR-Linac and assigned the designation of Volunteer 1 (20s-year-old male) and Volunteer 2 (20s-year-old female) for future comparisons. Additionally, a 50s-year-old male with American Joint Committee on Cancer (AJCC) 8^th^ Edition Stage I (cT2, cN0, cM0, p16+) human papilloma virus (HPV) positive of oropharynx, squamous cell carcinoma of the left tonsil was scanned on the MR-Sim to evaluate the potential of SyntheticMR in the radiation therapy workflow.

### 2.4. Data Collection and Image Processing

The images were processed using the SyntheticMR post-processing software, SyMRI (StandAlone 11.3.11, Linköping, Sweden) developed specifically for the 2D-MDME sequence. The geometric accuracy was evaluated using the ACR phantom by measuring four equal radially spaced diameters and comparing them to the expected length of 190 mm to within ±2 mm^34^. For the phantom analysis, a circular region-of-interest (ROI) was created in each vial using 3D Slicer^35^ (https://www.slicer.org/) using the synthetically reconstructed T2-weighted MRI and the values within each ROI were extracted for processing in the quantitative T1, T2, and PD maps. A margin of approximately 10% of the vial’s diameter was left to account for ringing artifacts hindering accurate readings^36^. For the *in vivo* analysis, the parotid and submandibular glands were chosen for analysis as the primary OAR for salivary dysfunction and were contoured automatically using a deep learning algorithm in the Advanced Medical Imaging Research Engine (ADMIRE) research software (v3.48.4, Elekta AB, Stockholm, Sweden). Additional critical structures including the tongue, bilateral infraorbital lymph spaces, mandible, and bilateral masseter muscles were contoured by medical students in RayStation Research 12A R v13.1.100.0 (RaySearch Laboratories, Stockholm, Sweden).

### 2.5. Statistical Analysis

All relevant analysis concerning statistical methods are formulated using the guidelines for reporting Statistical Analyses and Methods in the Published Literature (SAMPL)^37^. All computational analysis was completed using Python 3.8.10. To evaluate quantitative parameter accuracy, for each vial inside the ISMRM/NIST phantom, the values inside the ROI were extracted and the mean and standard deviation were calculated and compared to manufacturer reference values. For all calculations, analysis was restricted to clinical ranges of T1 (250 – 2000 ms), T2 (30 – 300 ms), and PD (20 – 160 pu)^38^ across both 1.5T and 3T MRI devices. Lin’s Concordance Correlation Coefficient (LCCC) was used instead of Pearson’s r^2^ to evaluate direct agreement to reference values instead of generalized linearity. For measuring bias between the measured values and reference values, the mean bias was determined, and Spearman’s rank correlation coefficient (i.e., Spearman’s ρ) was calculated to test for significant generalized correlations between bias and the magnitude of the reference value. A p-value of 0.05 was used for statistical significance. These trends were further visualized using a Bland-Altman plot^39^. Cluster analysis was performed using an elliptical envelope and a soft margin assuming 40% outliers. To assess the separation of the clusters for desired ROIs, the purity measure was used to describe the proportion of desired data points that are within the desired cluster compared to other ROIs. Statistical significance between desired ROIs was calculated using the SigClust method^40^ with soft thresholding and 100 Monte Carlo iterations following mean centering and variance normalization.

## 3. Results

### 3.1. Phantom Analysis

For the geometric accuracy / distortion analysis using the ACR large phantom, the MR-Sim and both sequences on the MR-Linac showed within-tolerance agreement with the 190 mm expected diameter for each of the four measurements. Further, lines drawn along the geometry grid lines were straight and showed no intra-phantom distortion.

When evaluating quantitative parameter accuracy, the Lin’s Concordance Correlation Coefficient between the measured and phantom reference values was above 0.97 for both the MR-Sim and MR-Linac when analyzing for T1, T2, and PD as shown in Figure 1. The highest agreement was seen in the PD values (average LCCC = 0.995), followed by T1 (average LCCC = 0.987), then T2 (average LCCC = 0.985). There is no significant difference between the LCCC of the MR-Sim and MR-Linac.

**Figure 1.**
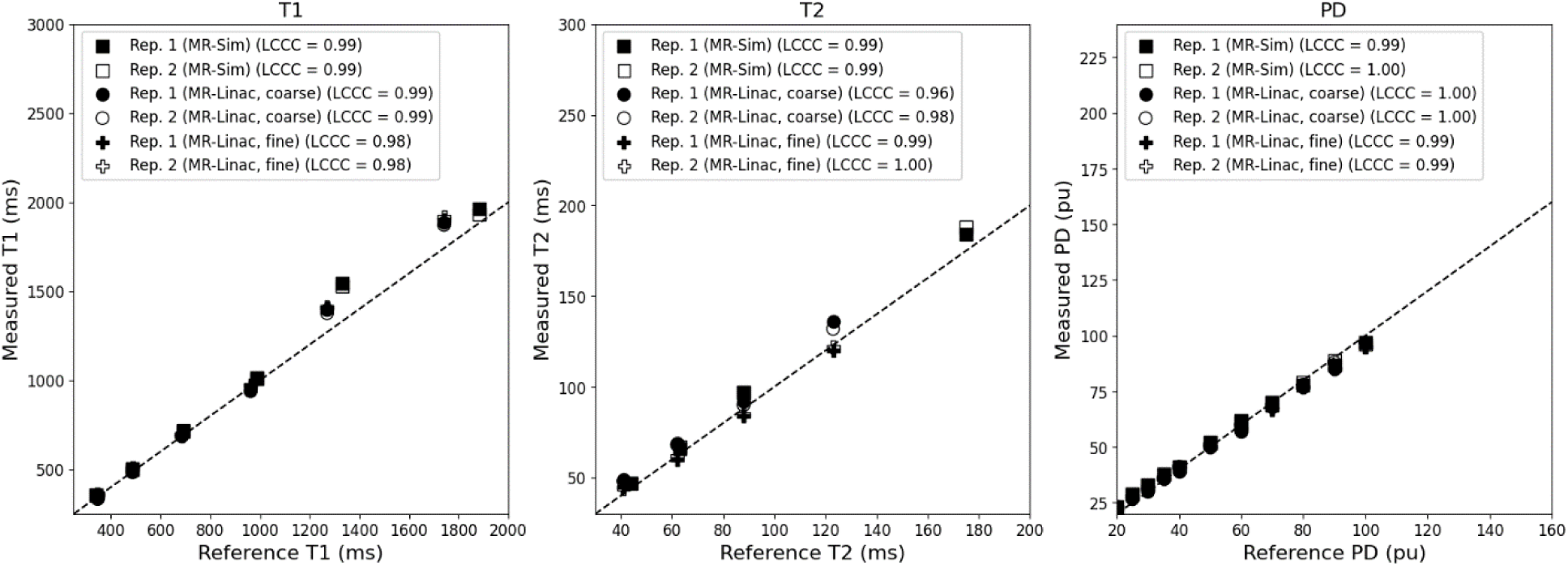
Correlation plot between the measured and reference T1 (left), T2 (center), and PD (right) for the MR-Sim and both coarse and fine sequences for the MR-Linac using a test-retest protocol. Abbreviations: LCCC = Lin’s Concordance Correlation Coefficient.

The mean bias between the measured and phantom reference values across both repeat scans was 4.22% for T1, 6.32% for T2, and 3.11% for PD for both the MR-Sim and MR-Linac as shown in Figure 2. The MR-Linac coarse sequence had the lowest average T1 bias at 0.19% while averaging 1.70% for T2 and 9.62% for PD. The MR-Linac fine sequence had the lowest average T2 and PD bias at 0.76& and 1.92%, respectively, while averaging 5.12% for T1. The MR-Sim averaged biases of 5.42% for T1, 7.35% for T2, and 5.62% for PD. When calculating the correlation between reference values and bias, only the PD values showed significant p-values (all p < 0.001) with a Spearman rank correlation coefficient averaging −0.96 across the MR-Sim and both sequences on the MR-Linac.

**Figure 2.**
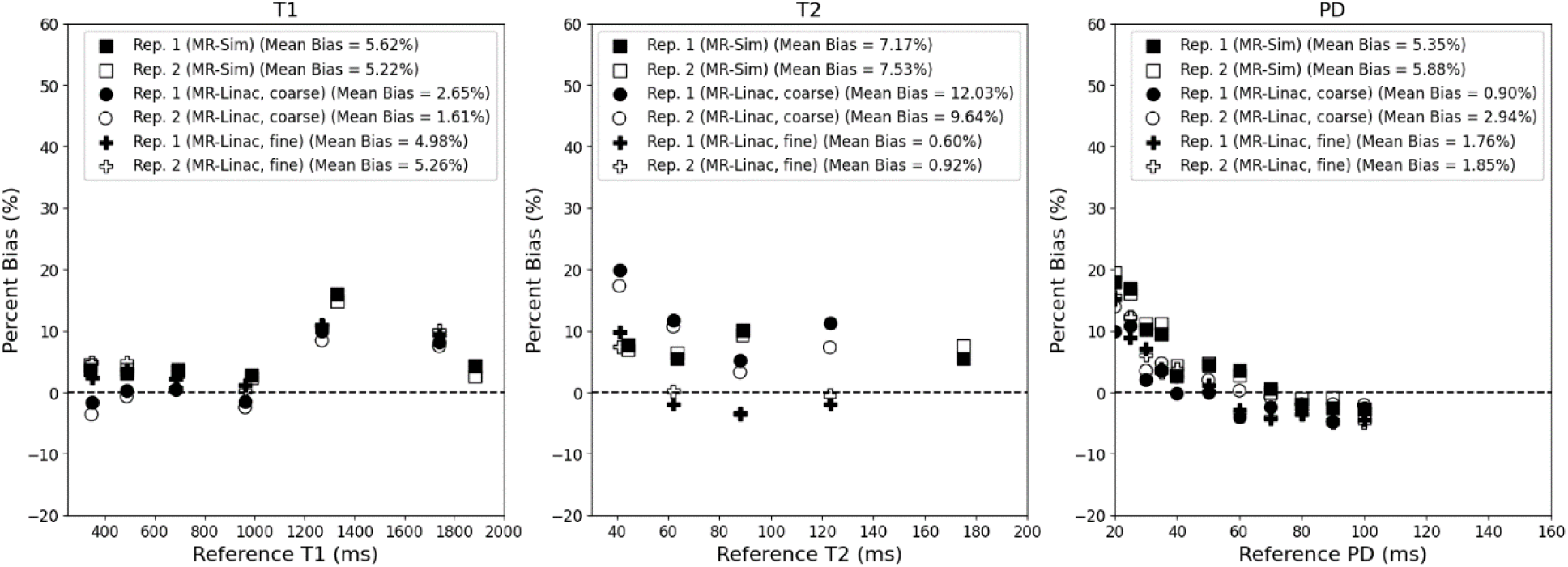
Bland-Altman plot between the measured and reference T1 (left), T2 (center), and PD (right) for the MR-Sim and both coarse and fine sequences for the MR-Linac using a test-retest protocol.

### 3.2. *In vivo* Analysis

An example of the SyMRI post-processing generated quantitative and subsequent synthetically generated contrast maps on the head and neck cancer patient on the MR-Sim in Figure 3. Note, the weighted contrast maps can be adjusted to different TE and TR values to achieve seemingly unlimited contrast options. Further, the inversion recovery (IR) sequences can be adjusted in a comparable way through the inversion time (TI).

**Figure 3.**
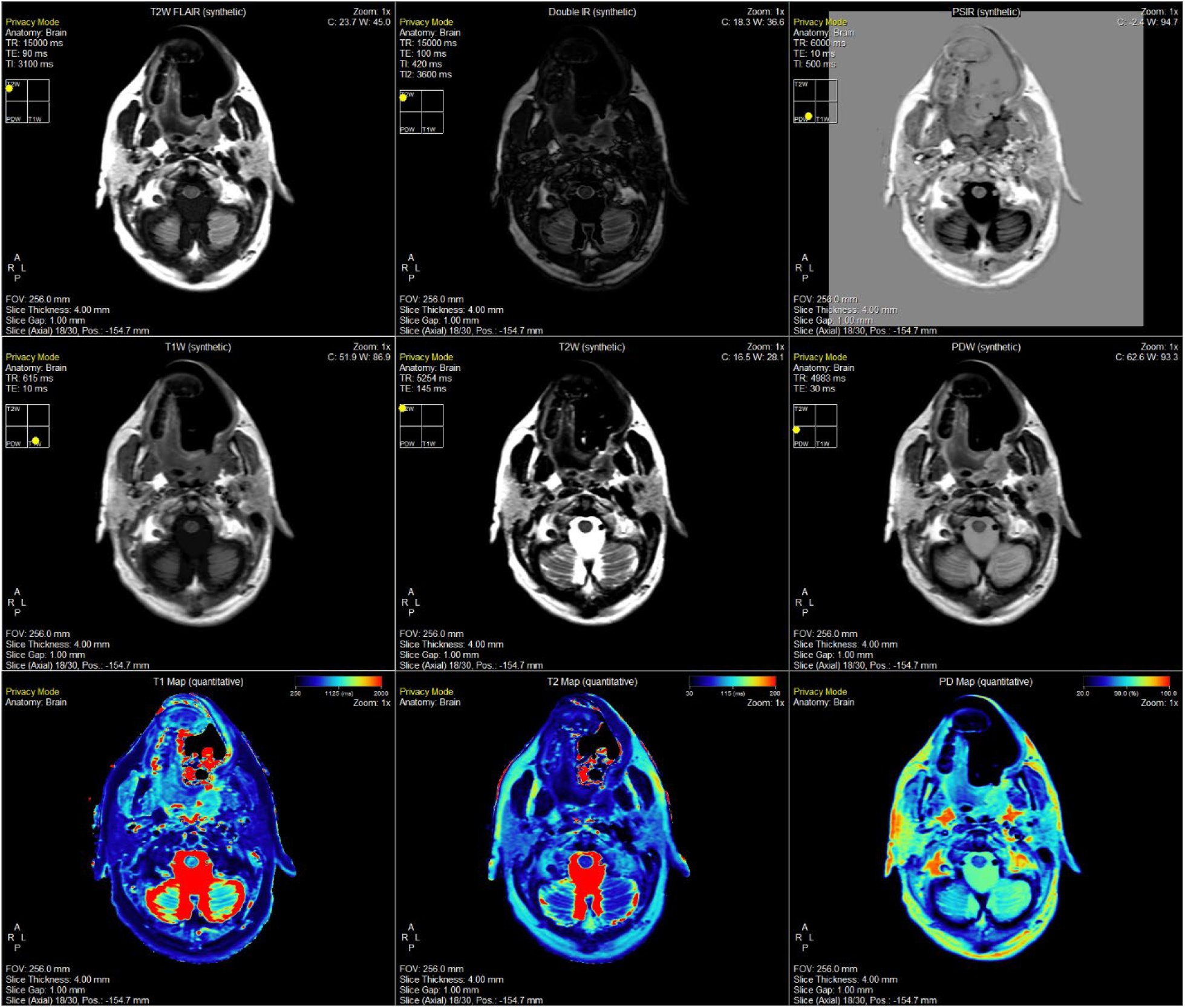
Demonstration of the SyMRI post-processing package offered by SyntheticMR for the head and neck cancer patient on the MR-Sim scanner. Shown here is an axial / transverse slice at the level of the parotid glands, medial pterygoid muscle, and masseter muscle. The same set of images are shown in the Supplementary Materials for Volunteer 1 on the MR-Sim (Figure 3-S1), Volunteer 2 on the MR-Sim (Figure 3-S2), Volunteer 1 on the MR-Linac coarse sequence (Figure 3-S3), and Volunteer 1 on the MR-Linac fine sequence (Figure 3-S4).

For the head and neck cancer patient, the T1, T2, and PD values inside each ROI were extracted and plotted against each other in two dimensions as shown in Figure 4. The purity measure was used between points inside each contour to determine cluster separability between the tumor and neighboring OARs. For T1 vs. T2 and T1 vs. PD, the GTV contour exhibited perfect purity while being 0.7 for T2 vs. PD. All cluster significance levels between the GTV and the nearest OAR, the tongue, using the SigClust method was p < 0.001.

**Figure 4.**
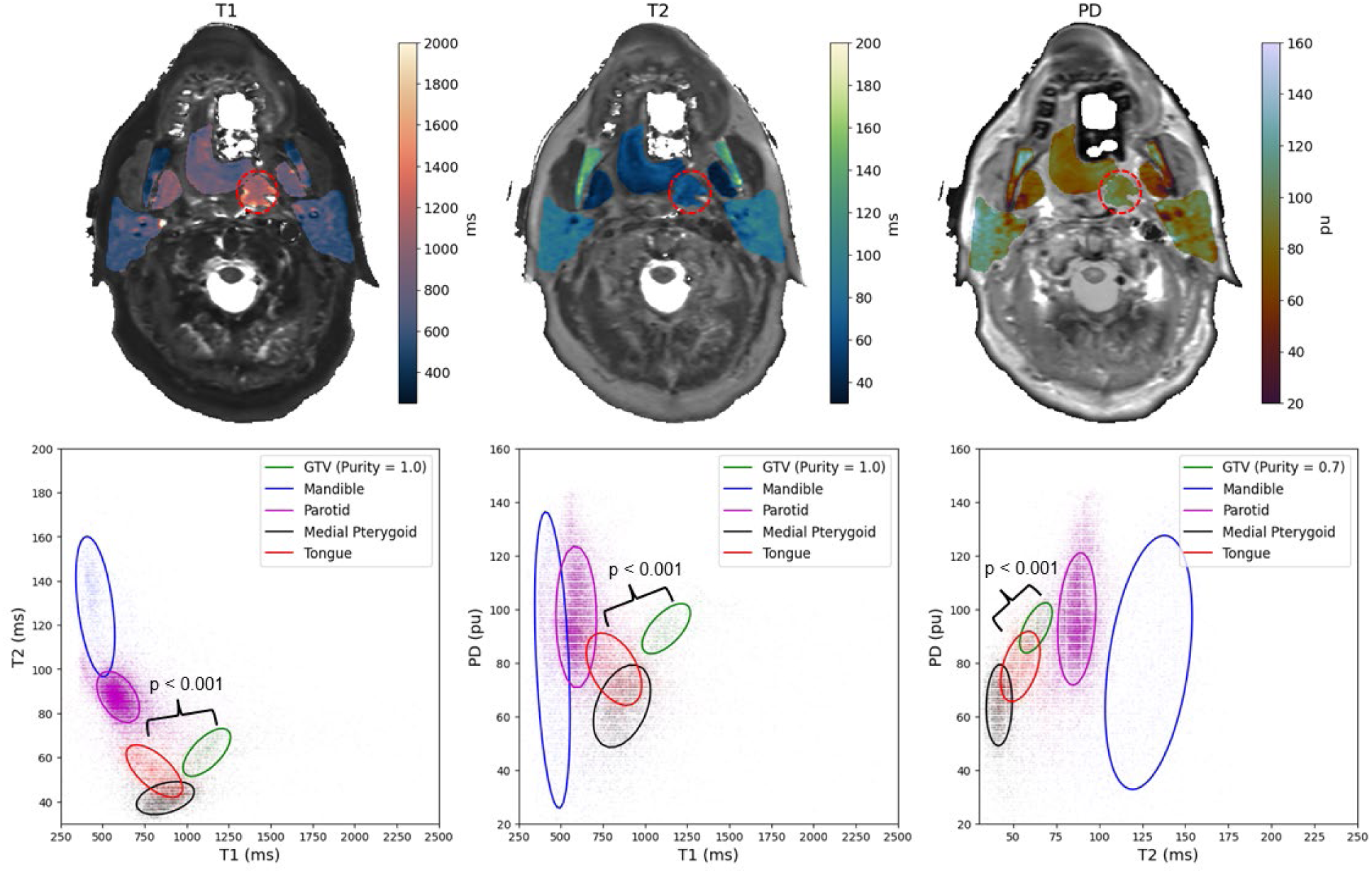
Above, the quantitative spatial maps of neighboring OARs for the patient on the MR-Sim overlayed on top of their respective synthetically generated contrast map. The gross tumor volume (GTV) is shown in red dashed circle. Note, this patient was wearing a bite block and a permanent retainer which caused significant signal dropout in the oral tongue as shown by the white regions. Below, a demonstration of the separation generated by SyntheticMR for differentiating tumor and neighboring healthy tissues. Additional quantitative cluster analysis for more additional OARs in healthy volunteers is shown in Figure 4-S1 of the Supplementary Materials.

## 4. Discussion

As shown in this technical feasibility analysis, SyntheticMR has the potential to create a paradigm shift in how imaging via MRI is done in the adaptive radiation oncology workflow. When comparing to a prior study collecting T1 and T2 measurements in parotid glands on 1.5T scanners^41^, the mean and standard deviation T1, 578.8 (67.9), and T2, 104.5 (11.7), agreed with the distribution shown in this paper. To the author’s knowledge, this is the first study investigating SyntheticMR on either the MR-Sim or MR-Linac. SyntheticMR can generate inherently co-registered T1, T2, and PD quantitative maps along with synthetically generated weighted and inversion recovery images in under six minutes. This has the potential to replace current time-intensive sequences and complex registration techniques currently used in the radiation oncology workflow for more efficient biomarker-based adaptive radiation therapy, increasing the adoption of more specialized quantitative imaging biomarkers approaches^42^ at high temporal density. Some previously studied examples in the head and neck which could be adopted due to the time savings of SyntheticMR include apparent diffusion coefficient (ADC)^43^, dynamic contrast enhanced (DCE)^44^, diffusion-weighted imaging (DWI)^45^, and more emerging biomarkers such as T1-rho^46^. Further, the geometric accuracy test was well within passing criteria providing sufficient confidence for radiation therapy setup, planning, and delivery.

Despite its promise, the current limitations of SyntheticMR include susceptibility to patient motion artifacts which would propagate to all generated maps, suppression of the blood signal causing black blood features, and ghosting artifacts due to flow sensitivity. Another simultaneous, multiparametric, technique which addresses some of these limitations and has been investigated by one prior group on the 1.5T MR-Linac^9^ is magnetic resonance fingerprinting, or MRF. This approach utilizes a series of dynamic scan acquisitions with each dynamic inheriting a pseudo-random set of acquisition parameters, typically the TR, TE, and flip angle. This pseudo-random acquired series is then matched to a pre-computed dictionary of MRI signal generation and fit to the most similar trajectory given the type of pulse sequence used. In comparison to SyntheticMR, MRF can acquire data using either a spin-echo or gradient-echo based approach allowing it to encode diffusion^47^and other weightings important in adaptive radiation therapy for head and neck cancer. However, MRF requires large pre-computed dictionary creation which may add prohibitive computational requirements. Further, MRF is only commercially available on Siemens MRI scanners providing only simultaneous T1 and T2 quantification at the time of this writing limiting its immediate clinical translation in comparison to SyntheticMR which is commercially available on Siemens, GE, and Philips.

**Figure 5:**
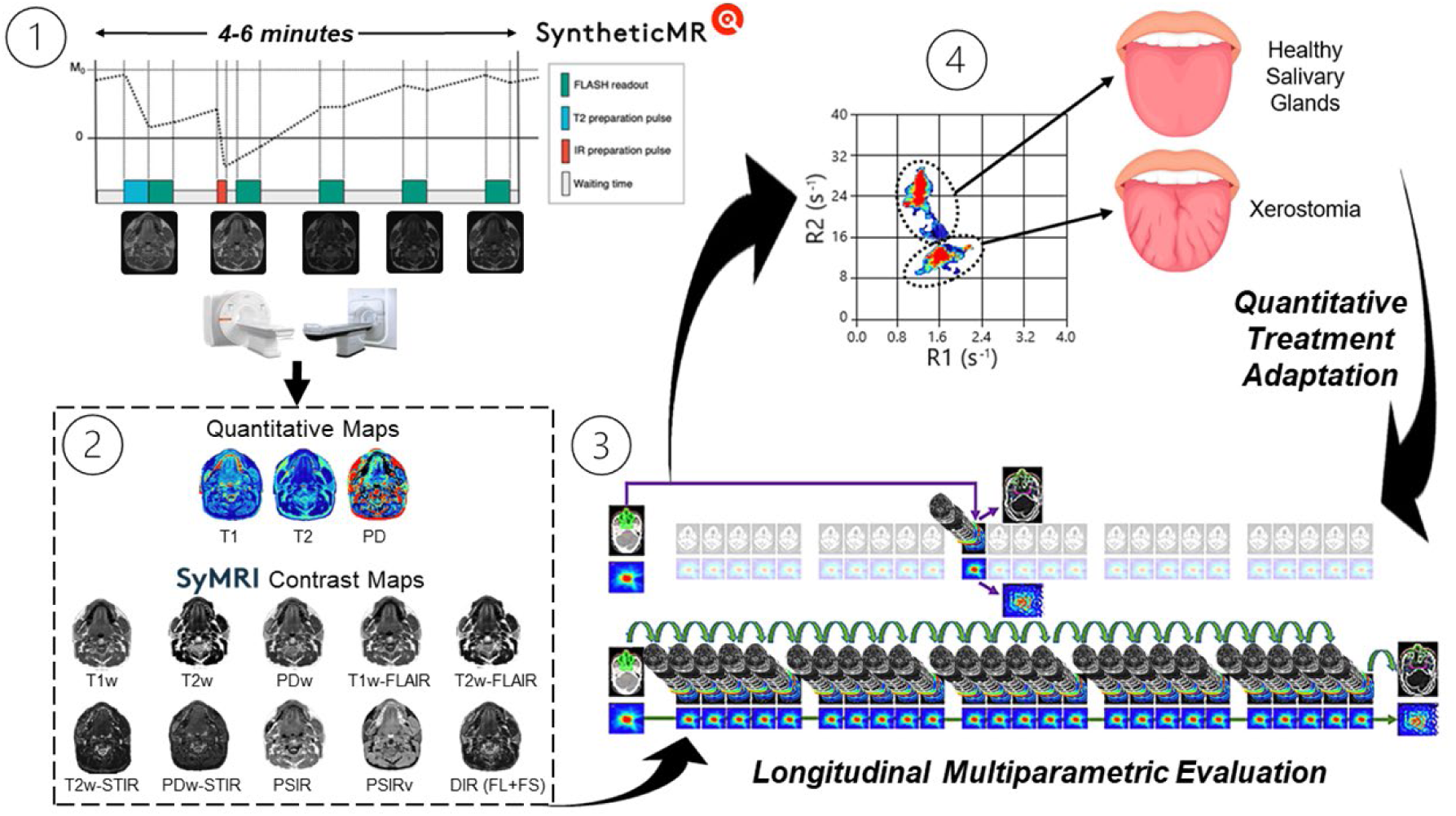
One potential application of SyntheticMR in the radiation oncology workflow to quantitatively adapt treatment based on detectable normal tissue damage in the salivary glands: 1) MRI acquisition overview, 2) generation of quantitative maps and subsequent synthetic contrast maps from SyMRI, 3) longitudinal acquisition schedule with high daily temporal resolution, and 4) subsequent evaluation of the deviations in the SyntheticMR maps. Note, the pulse sequence diagram (adapted from Fujita et al. 2024^15^) on the top left is for the 3D-QALAS sequence, not the 2D-MDME used in this study. Further, the figure on the bottom right is adapted from Heukelom and Fuller 2022^48^.

The future applications of SyntheticMR in the general radiation oncology workflow is wide. Recent movements have suggested the transition to probabilistic target definition instead of the currently used uniform target definitions^49^ which builds upon the ideas presented in dose painting^50^. SyntheticMR has the potential to become a valuable asset in this space due to its multiparametric quantitative input at the voxel level which will help to inform probabilistic target definitions and optimal dose painting strategies. Techniques such as Bayesian and spatial statistics may also be employed upon SyntheticMR output to assess treatment response, identify sub-volume boost regions, and create more robust tumor control probability (TCP) and normal tissue complication probability (NTCP) models.

The multiparametric quantitative input at the voxel level may also be used as additional input channels for advanced image segmentation algorithms. This approach may be done manually using spatial statistics approaches, or automatically using deep learning techniques to identify optimal boundaries for each desired ROI in the three-dimensional quantitative space (T1 vs. T2 vs. PD). These boundaries may be created using large cohort studies of healthy volunteers and those with malignancies to generate consensus clusters for each desired ROI. Additional features from the synthetically generated contrast maps, such as radiomics, may also be included for improved performance^24^. Similar research utilizing SyntheticMR to achieve these tasks in the brain for white matter, gray matter, and others has been successfully demonstrated and employed clinically as a product of SyntheticMR. Additionally, after these regions have been successfully determined and validated, they may be used to assess the error of clinical contours and quantify contour uncertainty in the presence of more homogeneous or heterogeneous ROIs.

## 5. Conclusion

SyntheticMR is a promising novel approach to consolidate the currently inhibitory time requirements for multiparametric MRI acquisition incorporating both anatomical scans and quantitative information. In this paper, we demonstrated the potential of SyntheticMR to enhance the radiation oncology workflow in the following ways: 1) achievement of multi-contrast anatomical information and quantification in a single scan acquisition time allowing for higher throughput or addition of more research sequences 2) superior simultaneous quantitative accuracy at higher spatial resolution compared to existing techniques, and 3) clinically acceptable repeatability, reproducibility, and spatial accuracy. These factors aligned with the high temporal resolution of the MR-Linac (i.e., 33 fractions for a head and neck cancer patient) will exponentially increase both the clinical and research efficiency of the MR-Linac inside the adaptive radiation therapy workflow as it is currently used allowing for more effective balancing of tumor control with reduction of normal tissue toxicity.

## Data Availability

All relevant anonymized SyntheticMR unprocessed (pre-SyMRI) DICOM files are to be made publicly available after manuscript acceptance at the following DOI:
10.6084/m9.figshare.26835715
The accompanying code for image visualization and statistical analysis will be made publicly available at the following URL:
https://github.com/Lucas-Mc/SyntheticMR_R-IDEAL_0-1

https://doi.org/10.6084/m9.figshare.26835715

https://github.com/Lucas-Mc/SyntheticMR_R-IDEAL_0-1.git

## Contributor Roles Taxonomy (CRediT) Attribution Statement

*Conceptualization*: L.M. and C.D.F.; *Data curation*: L.M. and K.H.; *Formal analysis*: L.M.; *Funding acquisition*: C.D.F.; *Investigation*: L.M., S.J.M., N.W. and T.C.S.; *Methodology*: L.M. and C.D.F.; *Project administration*: C.D.F.; *Resources*: L.M., S.J.M., N.W., R.A., A.M., H.S., T.C.S., Y.D., E.S., K.H. and C.D.F.; *Software*: L.M.; *Supervision*: A.D., E.S., D.M., R.J.S., K.H. and C.D.F.; *Validation*: L.M., S.J.M. and N.W.; *Visualization*: L.M.; *Writing – original draft*: L.M.; *Writing - review & editing*: L.M., S.J.M., N.W., R.A., A.M., H.S., T.C.S., Y.D., A.D., E.S., D.M., R.J.S., K.H. and C.D.F.

## IRB Statement

All participants provided written informed consent. Volunteers were consented to an internal volunteer imaging protocol (PA15-0418), both approved by the institutional review board at The University of Texas MD Anderson Cancer Center.

## Data Availability Statement

All relevant anonymized SyntheticMR unprocessed (pre-SyMRI) DICOM files are to be made publicly available after manuscript acceptance at the following DOI: 10.6084/m9.figshare.26835715. The accompanying code for image visualization and statistical analysis will be made publicly available at the following URL: https://github.com/Lucas-Mc/SyntheticMR_R-IDEAL_0-1.

## Acknowledgements

The authors would like to thank SyntheticMR AB for their assistance in the formal data analysis through their software, SyMRI.

---

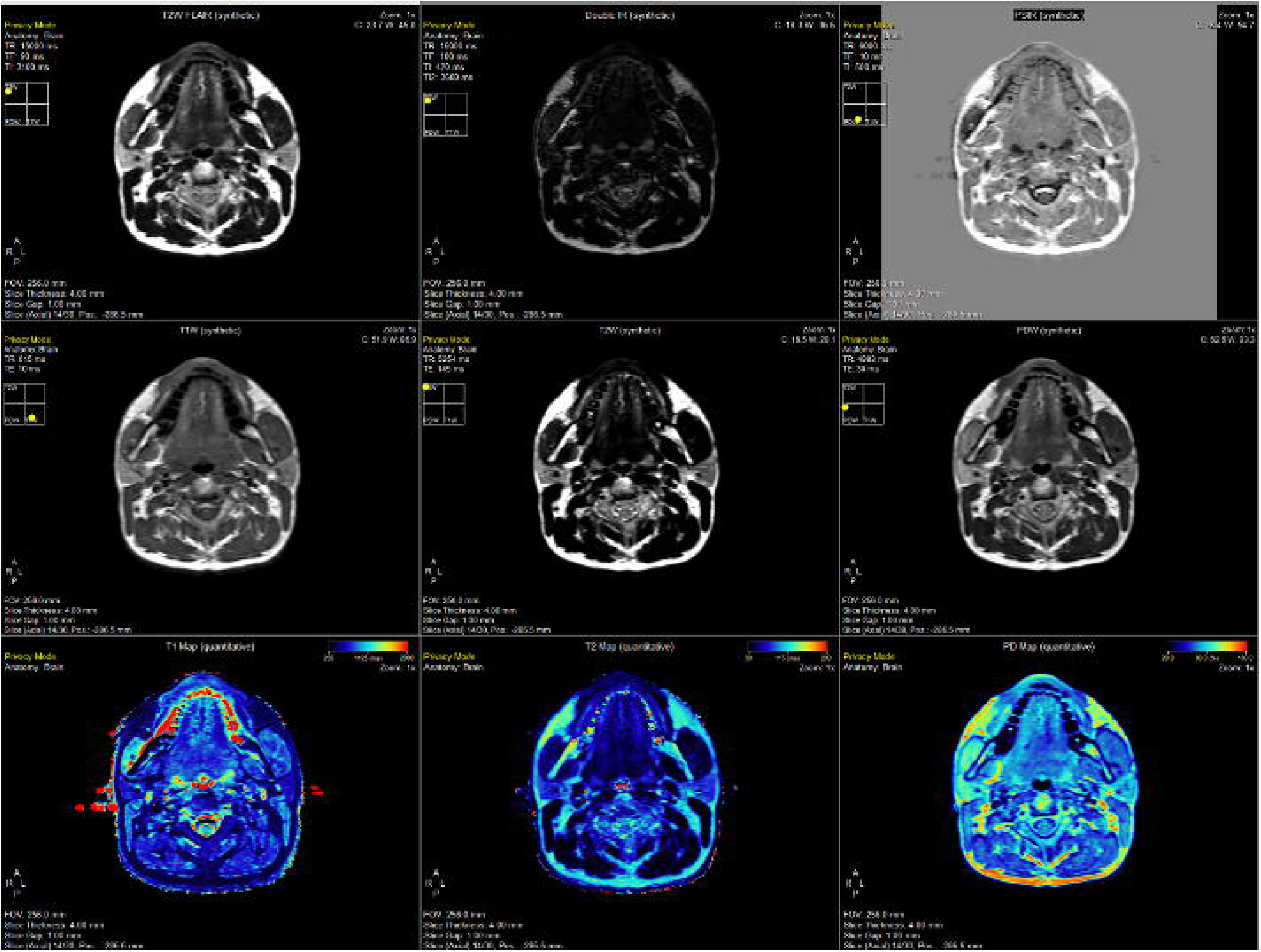

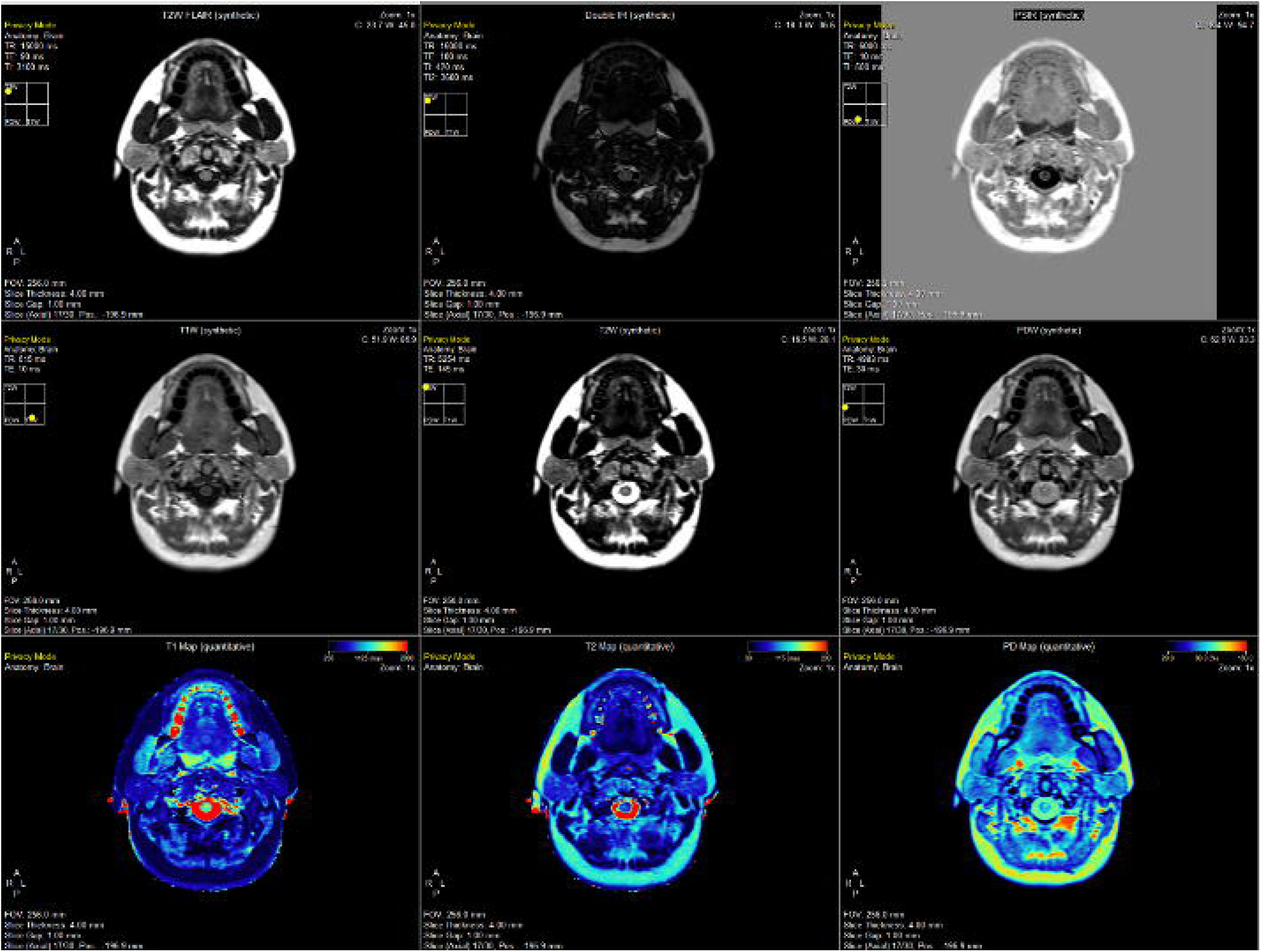

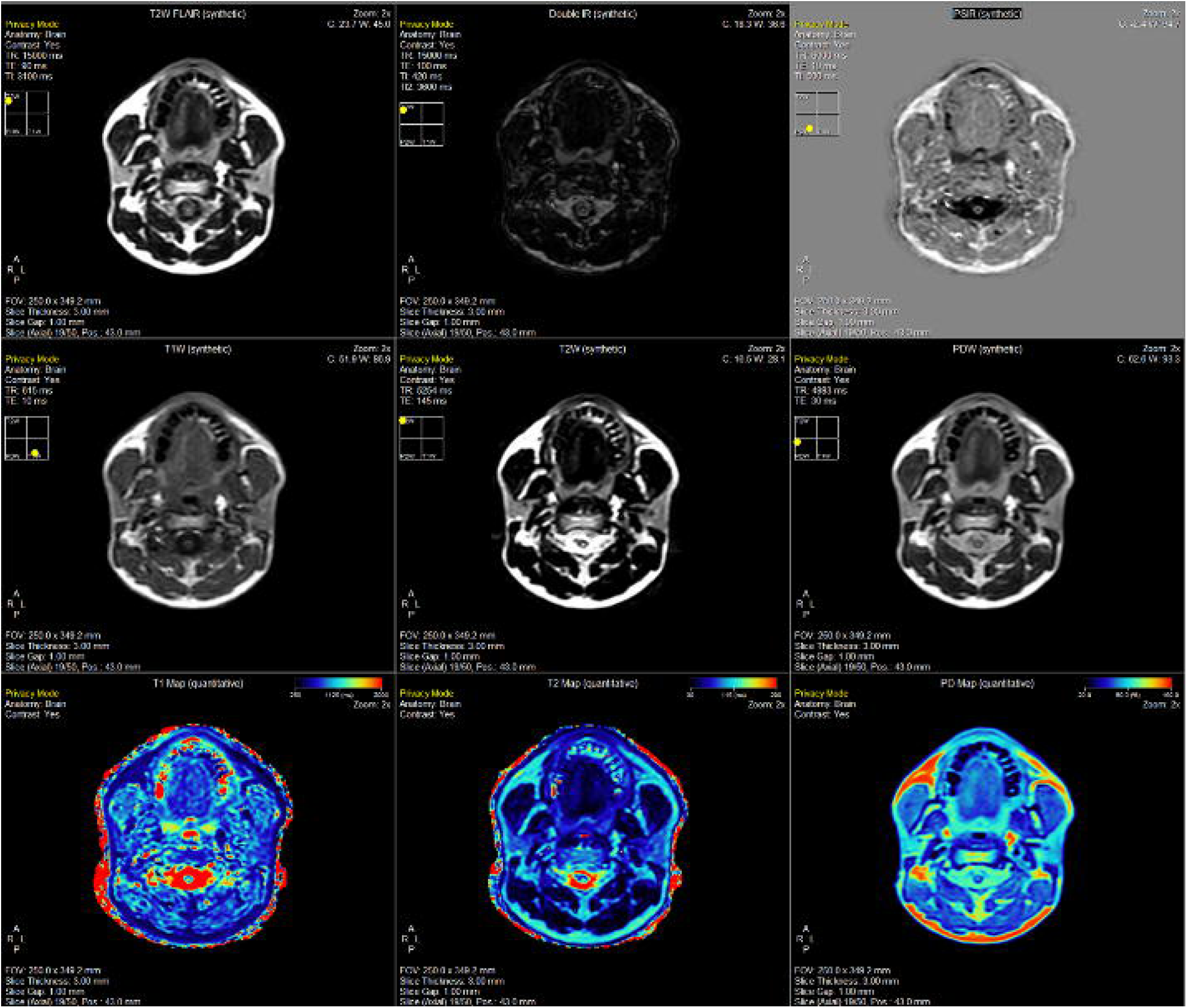

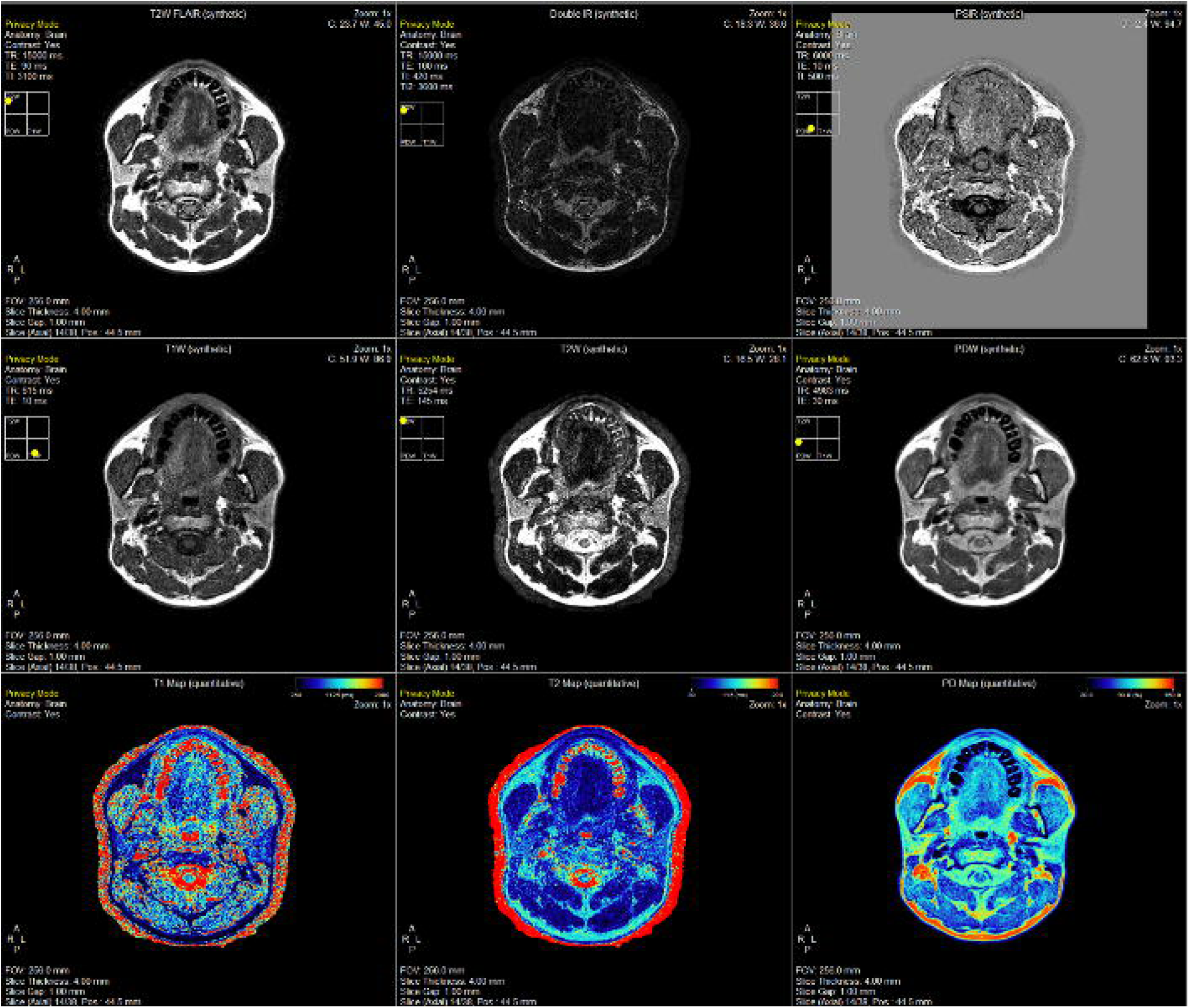

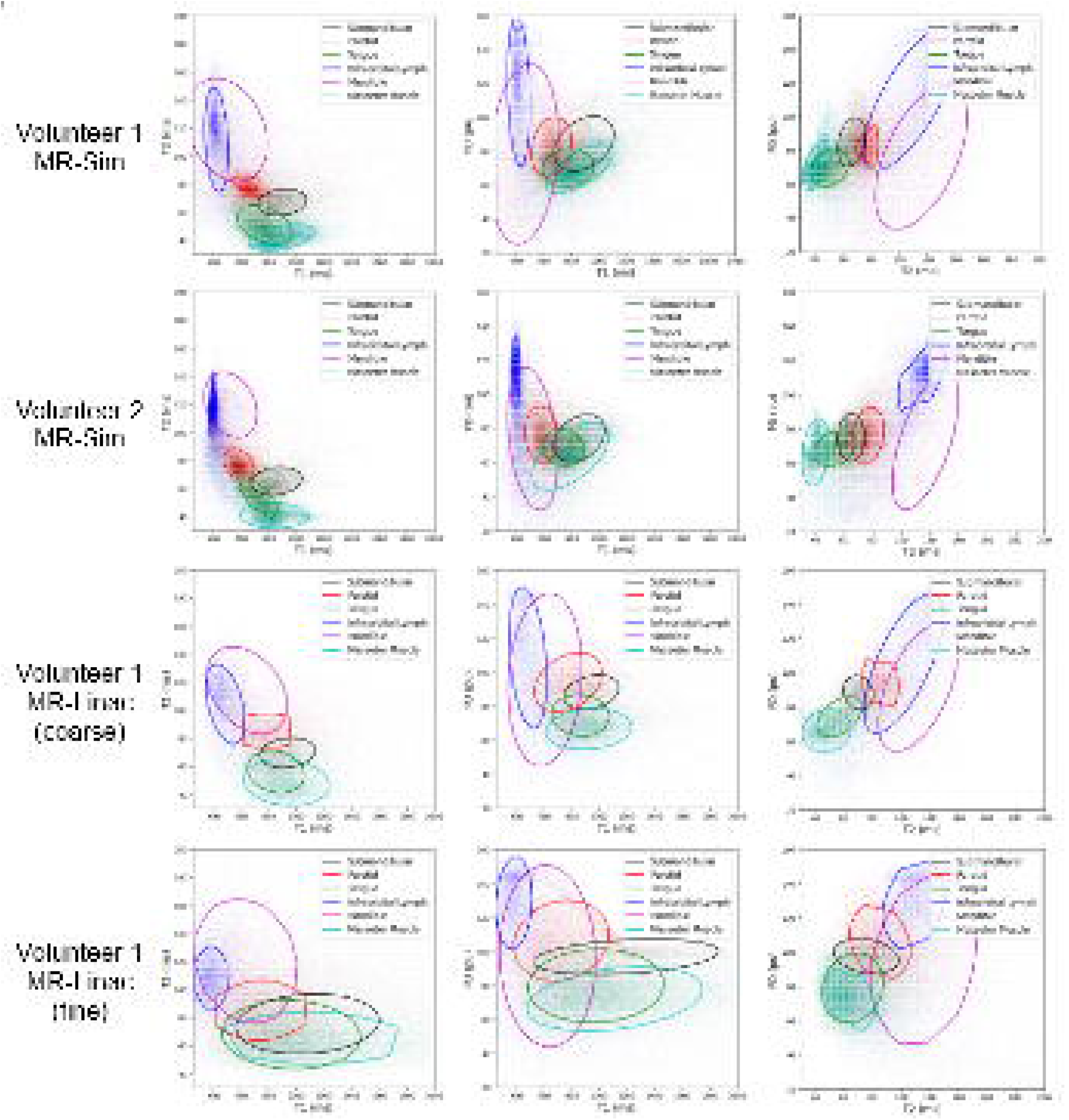

## References

1. Shankar L, Montanera W. Computed Tomography Versus Magnetic Resonance Imaging and Three-Dimensional Applications. Medical Clinics of North America. 1991;75(6):1355–1366. doi:10.1016/S0025-7125(16)30392-3

2. Raaymakers BW, Lagendijk JJW, Overweg J, et al. Integrating a 1.5 T MRI scanner with a 6 MV accelerator: proof of concept. Phys Med Biol. 2009;54(12):N229–N237. doi:10.1088/0031-9155/54/12/N01

3. Cashmore MT, McCann AJ, Wastling SJ, McGrath C, Thornton J, Hall MG. Clinical quantitative MRI and the need for metrology. The British Journal of Radiology. 2021;94(1120):20201215. doi:10.1259/bjr.20201215

4. Qu J, Pan B, Su T, et al. T1 and T2 mapping for identifying malignant lymph nodes in head and neck squamous cell carcinoma. Cancer Imaging. 2023;23(1):125. doi:10.1186/s40644-023-00648-6

5. Kim SE, Roberts JA, Kholmovski EG, Hitchcock Y, Anzai Y. T1 mapping for Head and Neck Cancer Patients undergoing Chemoradiotherapy: Feasibility of 3D Stack of Star Imaging. Magnetic Resonance Imaging. 2024;112:38–46. doi:10.1016/j.mri.2024.04.005

6. Guerreiro F, Van Houdt PJ, Navest RJM, et al. Validation of quantitative magnetic resonance imaging techniques in head and neck healthy structures involved in the salivary and swallowing function: Accuracy and repeatability. Physics and Imaging in Radiation Oncology. 2024;31:100608. doi:10.1016/j.phro.2024.100608

7. Kooreman ES, Van Houdt PJ, Nowee ME, et al. Feasibility and accuracy of quantitative imaging on a 1.5 T MR-linear accelerator. Radiotherapy and Oncology. 2019;133:156–162. doi:10.1016/j.radonc.2019.01.011

8. Subashi E, LoCastro E, Apte A, Zelefsky MJ, Tyagi N. Quantitative Relaxometry for Target Localization and Response Assessment in Ultra-Hypofractionated MR-Guided Radiotherapy to the Prostate and DIL. International Journal of Radiation Oncology*Biology*Physics. 2022;114(3):S33. doi:10.1016/j.ijrobp.2022.07.390

9. Bruijnen T, Van Der Heide O, Intven MPW, et al. Technical feasibility of magnetic resonance fingerprinting on a 1.5T MRI-linac. Phys Med Biol. 2020;65(22):22NT01. doi:10.1088/1361-6560/abbb9d

10. Grimbergen G, Hackett SL, Van Ommen F, et al. Gating and intrafraction drift correction on a 1.5 T MR-Linac: Clinical dosimetric benefits for upper abdominal tumors. Radiotherapy and Oncology. 2023;189:109932. doi:10.1016/j.radonc.2023.109932

11. Warntjes JBM, Leinhard OD, West J, Lundberg P. Rapid magnetic resonance quantification on the brain: Optimization for clinical usage. Magnetic Resonance in Med. 2008;60(2):320–329. doi:10.1002/mrm.21635

12. Hagiwara A, Hori M, Cohen-Adad J, et al. Linearity, Bias, Intrascanner Repeatability, and Interscanner Reproducibility of Quantitative Multidynamic Multiecho Sequence for Rapid Simultaneous Relaxometry at 3 T: A Validation Study With a Standardized Phantom and Healthy Controls. Invest Radiol. 2019;54(1):39–47. doi:10.1097/RLI.0000000000000510

13. Hagiwara A, Warntjes M, Hori M, et al. SyMRI of the Brain: Rapid Quantification of Relaxation Rates and Proton Density, With Synthetic MRI, Automatic Brain Segmentation, and Myelin Measurement. Invest Radiol. 2017;52(10):647–657. doi:10.1097/RLI.0000000000000365

14. Kvernby S, Warntjes MJB, Haraldsson H, Carlhäll CJ, Engvall J, Ebbers T. Simultaneous three-dimensional myocardial T1 and T2 mapping in one breath hold with 3D-QALAS. Journal of Cardiovascular Magnetic Resonance. 2014;16(1):102. doi:10.1186/s12968-014-0102-0

15. Fujita S, Hagiwara A, Hori M, et al. Three-dimensional high-resolution simultaneous quantitative mapping of the whole brain with 3D-QALAS: An accuracy and repeatability study. Magnetic Resonance Imaging. 2019;63:235–243. doi:10.1016/j.mri.2019.08.031

16. Fujita S, Gagoski B, Hwang K, et al. Cross-vendor multiparametric mapping of the human brain using 3D-QALAS : A multicenter and multivendor study. Magnetic Resonance in Med. 2024;91(5):1863–1875. doi:10.1002/mrm.29939

17. Hwang K, Fujita S. Synthetic MR: Physical principles, clinical implementation, and new developments. Medical Physics. 2022;49(7):4861–4874. doi:10.1002/mp.15686

18. Xu S, Ma Z, Zhang J, et al. Quantitative assessment of preoperative brain development in pediatric congenital heart disease patients by synthetic MRI. Insights Imaging. 2024;15(1):166. doi:10.1186/s13244-024-01746-0

19. Zhang P, Yang J, Shu Y, et al. The value of synthetic MRI in detecting the brain changes and hearing impairment of children with sensorineural hearing loss. Front Neurosci. 2024;18:1365141. doi:10.3389/fnins.2024.1365141

20. Lin L, Chen Y, Dai Y, et al. Quantification of myelination in children with attention-deficit/hyperactivity disorder: a comparative assessment with synthetic MRI and DTI. Eur Child Adolesc Psychiatry. 2024;33(6):1935–1944. doi:10.1007/s00787-023-02297-3

21. Zhang L, Mai W, Mo X, et al. Quantitative evaluation of meniscus injury using synthetic magnetic resonance imaging. BMC Musculoskelet Disord. 2024;25(1):292. doi:10.1186/s12891-024-07375-4

22. Qu M, Feng W, Liu X, et al. Investigation of synthetic MRI with quantitative parameters for discriminating axillary lymph nodes status in invasive breast cancer. European Journal of Radiology. 2024;175:111452. doi:10.1016/j.ejrad.2024.111452

23. Chen Y, Meng T, Cao W, et al. Histogram analysis of MR quantitative parameters: are they correlated with prognostic factors in prostate cancer? Abdom Radiol. 2024;49(5):1534–1544. doi:10.1007/s00261-024-04227-6

24. Hwang KP, Elshafeey NA, Kotrotsou A, et al. A Radiomics Model Based on Synthetic MRI Acquisition for Predicting Neoadjuvant Systemic Treatment Response in Triple-Negative Breast Cancer. Radiology: Imaging Cancer. 2023;5(4):e230009. doi:10.1148/rycan.230009

25. Ljusberg A, Blystad I, Lundberg P, Adolfsson E, Tisell A. Radiation-dependent demyelination in normal appearing white matter in glioma patients, determined using quantitative magnetic resonance imaging. Physics and Imaging in Radiation Oncology. 2023;27:100451. doi:10.1016/j.phro.2023.100451

26. Zhang H, Hu L, Qin F, et al. Synthetic MRI and diffusion-weighted imaging for differentiating nasopharyngeal lymphoma from nasopharyngeal carcinoma: combination with morphological features. British Journal of Radiology. 2024;97(1159):1278-1285. doi:10.1093/bjr/tqae095

27. Konar AS, Paudyal R, Shah AD, et al. Qualitative and Quantitative Performance of Magnetic Resonance Image Compilation (MAGiC) Method: An Exploratory Analysis for Head and Neck Imaging. Cancers. 2022;14(15):3624. doi:10.3390/cancers14153624

28. Verkooijen HM, Kerkmeijer LGW, Fuller CD, et al. R-IDEAL: A Framework for Systematic Clinical Evaluation of Technical Innovations in Radiation Oncology. Front Oncol. 2017;7. doi:10.3389/fonc.2017.00059

29. Joint Head and Neck Radiotherapy-MRI Development Cooperative, Mohamed ASR, Abusaif A, et al. Prospective validation of diffusion-weighted MRI as a biomarker of tumor response and oncologic outcomes in head and neck cancer: Results from an observational biomarker pre-qualification study. Published online April 18, 2022. doi:10.1101/2022.04.18.22273782

30. Benedict SH, Yenice KM, Followill D, et al. Stereotactic body radiation therapy: The report of AAPM Task Group 101. Medical Physics. 2010;37(8):4078–4101. doi:10.1118/1.3438081

31. Vishwanath V, Jafarieh S, Rembielak A. The role of imaging in head and neck cancer:An overview of different imaging modalitiesin primary diagnosis and staging of the disease. jcb. 2020;12(5):512–518. doi:10.5114/jcb.2020.100386

32. Brahmbhatt S, Overfield CJ, Rhyner PA, Bhatt AA. Imaging of the Posttreatment Head and Neck: Expected Findings and Potential Complications. Radiology: Imaging Cancer. 2024;6(1):e230155. doi:10.1148/rycan.230155

33. Carr ME, Keenan KE, Rai R, Metcalfe P, Walker A, Holloway L. Determining the longitudinal accuracy and reproducibility of T _1_ and T _2_ in a 3T MRI scanner. J Applied Clin Med Phys. 2021;22(11):143–150. doi:10.1002/acm2.13432

34. Tijssen RHN, Philippens MEP, Paulson ES, et al. MRI commissioning of 1.5T MR-linac systems – a multi-institutional study. Radiotherapy and Oncology. 2019;132:114–120. doi:10.1016/j.radonc.2018.12.011

35. Fedorov A, Beichel R, Kalpathy-Cramer J, et al. 3D Slicer as an image computing platform for the Quantitative Imaging Network. Magnetic Resonance Imaging. 2012;30(9):1323–1341. doi:10.1016/j.mri.2012.05.001

36. Stupic KF, Ainslie M, Boss MA, et al. A standard system phantom for magnetic resonance imaging. Magnetic Resonance in Med. 2021;86(3):1194–1211. doi:10.1002/mrm.28779

37. Lang TA, Altman DG. Basic statistical reporting for articles published in Biomedical Journals: The “Statistical Analyses and Methods in the Published Literature” or the SAMPL Guidelines. International Journal of Nursing Studies. 2015;52(1):5–9. doi:10.1016/j.ijnurstu.2014.09.006

38. Bojorquez JZ, Bricq S, Acquitter C, Brunotte F, Walker PM, Lalande A. What are normal relaxation times of tissues at 3 T? Magnetic Resonance Imaging. 2017;35:69–80. doi:10.1016/j.mri.2016.08.021

39. Altman DG, Bland JM. Measurement in Medicine: The Analysis of Method Comparison Studies. The Statistician. 1983;32(3):307. doi:10.2307/2987937

40. Liu Y, Hayes DN, Nobel A, Marron JS. Statistical Significance of Clustering for High-Dimension, Low–Sample Size Data. Journal of the American Statistical Association. 2008;103(483):1281–1293. doi:10.1198/016214508000000454

41. Liao J, Saito N, Ozonoff A, Jara H, Steinberg M, Sakai O. Quantitative MRI analysis of salivary glands in sickle cell disease. Dentomaxillofac Radiol. 2012;41(8):630–636. doi:10.1259/dmfr/31672000

42. Van Houdt PJ, Saeed H, Thorwarth D, et al. Integration of quantitative imaging biomarkers in clinical trials for MR-guided radiotherapy: Conceptual guidance for multicentre studies from the MR-Linac Consortium Imaging Biomarker Working Group. European Journal of Cancer. 2021;153:64–71. doi:10.1016/j.ejca.2021.04.041

43. El-Habashy DM, Wahid KA, He R, et al. Longitudinal diffusion and volumetric kinetics of head and neck cancer magnetic resonance on a 1.5 T MR-linear accelerator hybrid system: A prospective R-IDEAL stage 2a imaging biomarker characterization/pre-qualification study. Clinical and Translational Radiation Oncology. 2023;42:100666. doi:10.1016/j.ctro.2023.100666

44. Khamis Y, Mohamed AS, Abobakr M, et al. Dynamic Contrast Enhanced MRI as a Biomarker of Tumor Response and Oncologic Outcomes in Head and Neck Cancer: Results of a Single Institution Prospective Imaging Study. International Journal of Radiation Oncology*Biology*Physics. 2023;117(2):e677–e678. doi:10.1016/j.ijrobp.2023.06.2134

45. McDonald BA, Salzillo T, Mulder S, et al. Prospective evaluation of in vivo and phantom repeatability and reproducibility of diffusion-weighted MRI sequences on 1.5 T MRI-linear accelerator (MR-Linac) and MR simulator devices for head and neck cancers. Radiotherapy and Oncology. 2023;185:109717. doi:10.1016/j.radonc.2023.109717

46. Kooreman ES, Tanaka M, Ter Beek LC, et al. T1ρ for Radiotherapy Treatment Response Monitoring in Rectal Cancer Patients: A Pilot Study. JCM. 2022;11(7):1998. doi:10.3390/jcm11071998

47. Afzali M, Mueller L, Sakaie K, et al. MR Fingerprinting with b-Tensor Encoding for Simultaneous Quantification of Relaxation and Diffusion in a Single Scan. Magnetic Resonance in Med. 2022;88(5):2043–2057. doi:10.1002/mrm.29352

48. Heukelom J, Fuller CD. Head and Neck Cancer Adaptive Radiation Therapy (ART): Conceptual Considerations for the Informed Clinician. Seminars in Radiation Oncology. 2019;29(3):258–273. doi:10.1016/j.semradonc.2019.02.008

49. Ferjančič P, Van Der Heide UA, Ménard C, Jeraj R. Probabilistic target definition and planning in patients with prostate cancer. Phys Med Biol. 2021;66(21):215011. doi:10.1088/1361-6560/ac2f8a

50. Fredén E, Tilly D, Ahnesjö A. Adaptive dose painting for prostate cancer. Front Oncol. 2022;12:973067. doi:10.3389/fonc.2022.973067

