## Supplementary Materials for "Technical Development and In Silico Implementation of SyntheticMR in Head and Neck Adaptive Radiation Therapy: A Prospective R-IDEAL Stage 0/1 Technology Development Report"

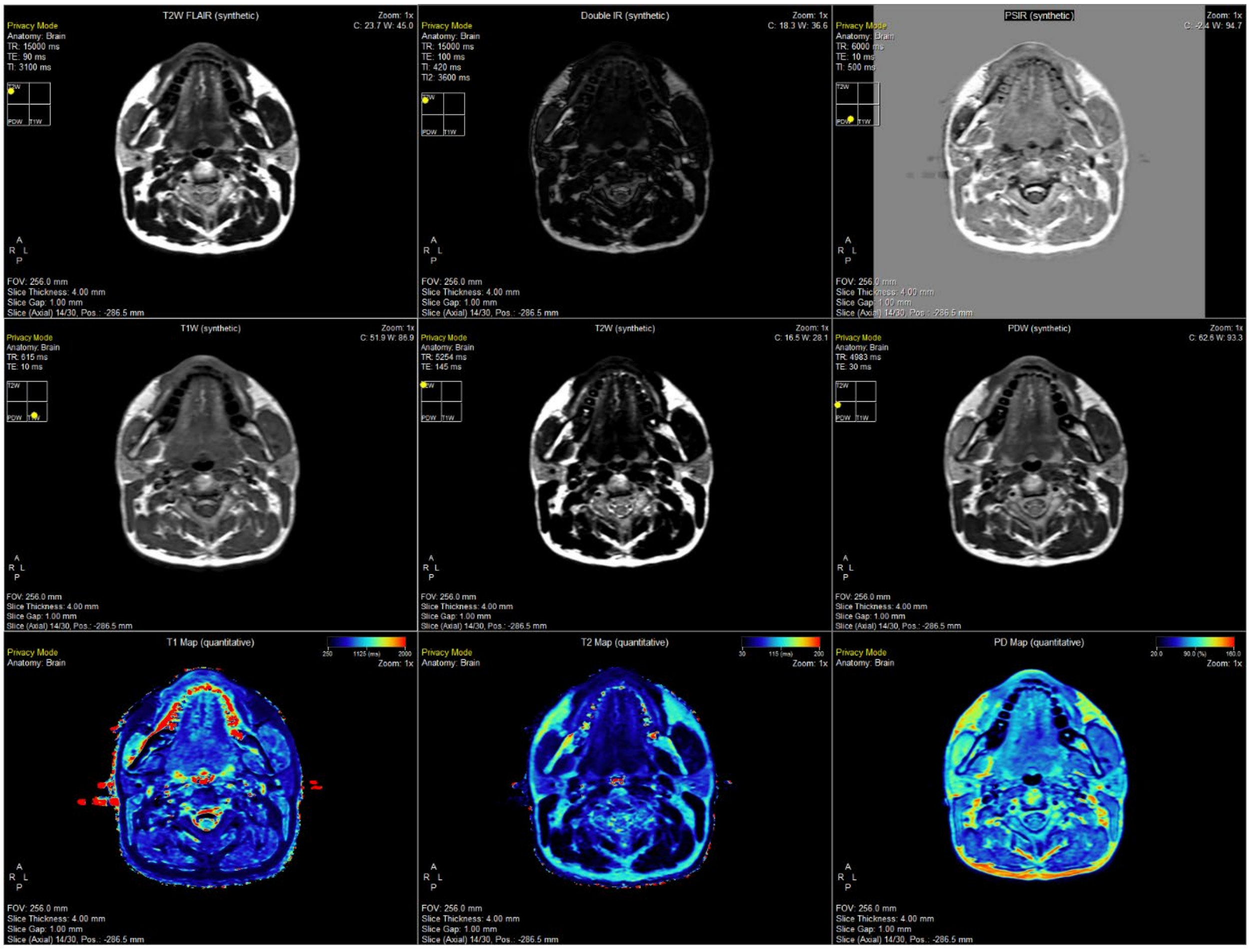


Figure 3-S1. Demonstration of the SyMRI post-processing package offered by SyntheticMR in Volunteer 1 on the MR-Sim scanner.

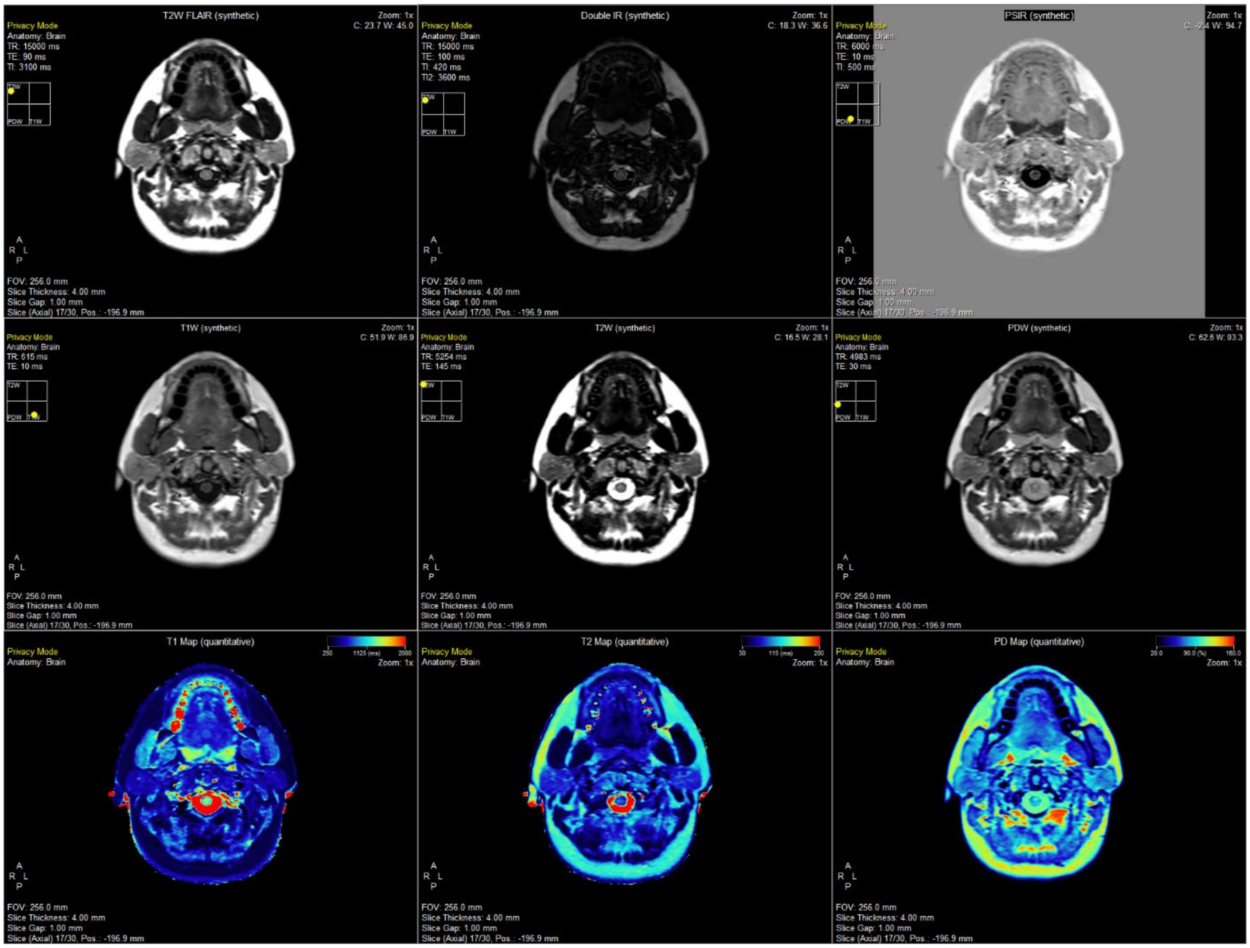

Figure 3-S2. Demonstration of the SyMRI post-processing package offered by SyntheticMR in Volunteer 2 on the MR-Sim scanner.

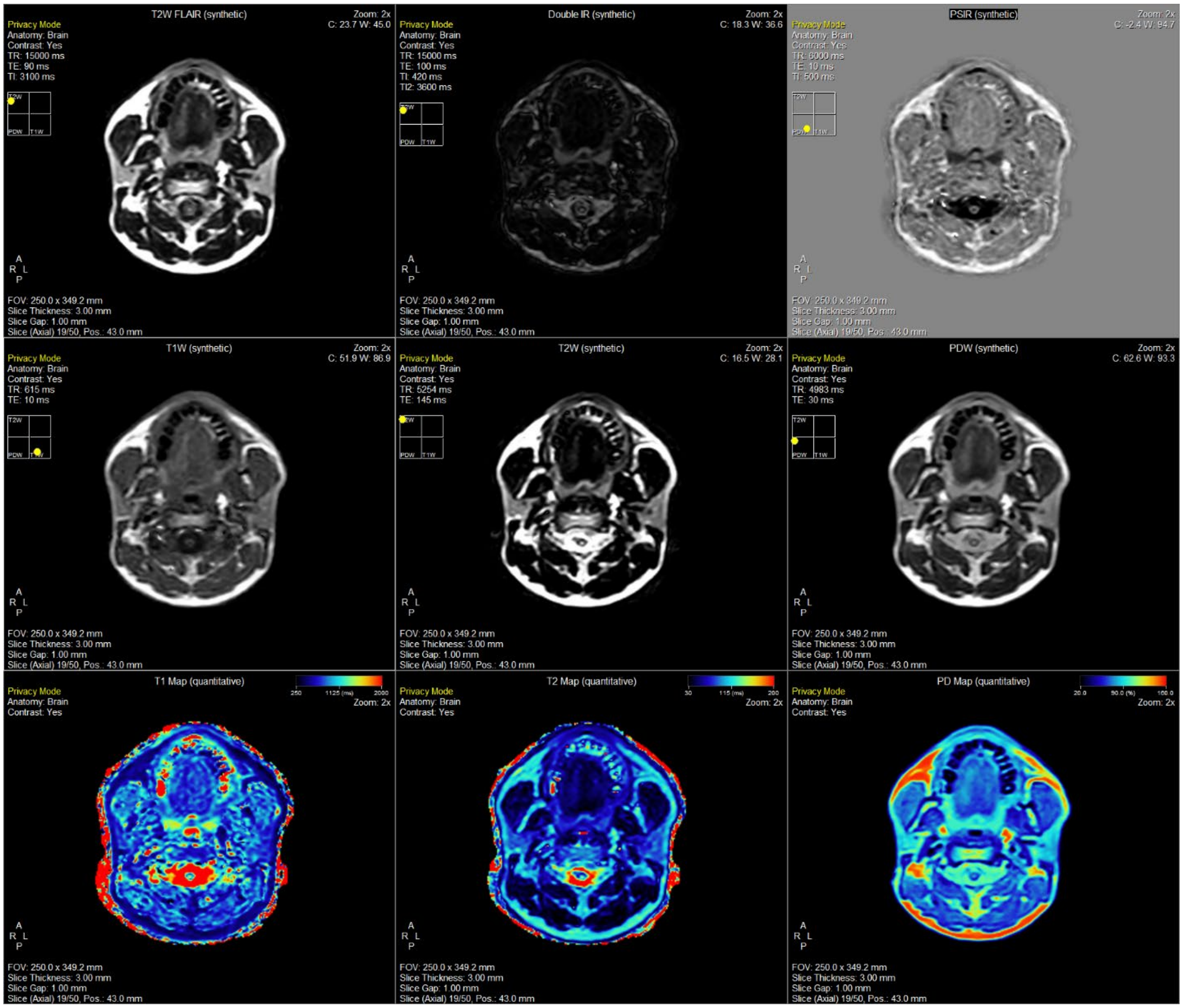

Figure 3-S3. Demonstration of the SyMRI post-processing package offered by SyntheticMR in Volunteer 1 using the coarse sequence on the MR-Linac scanner.

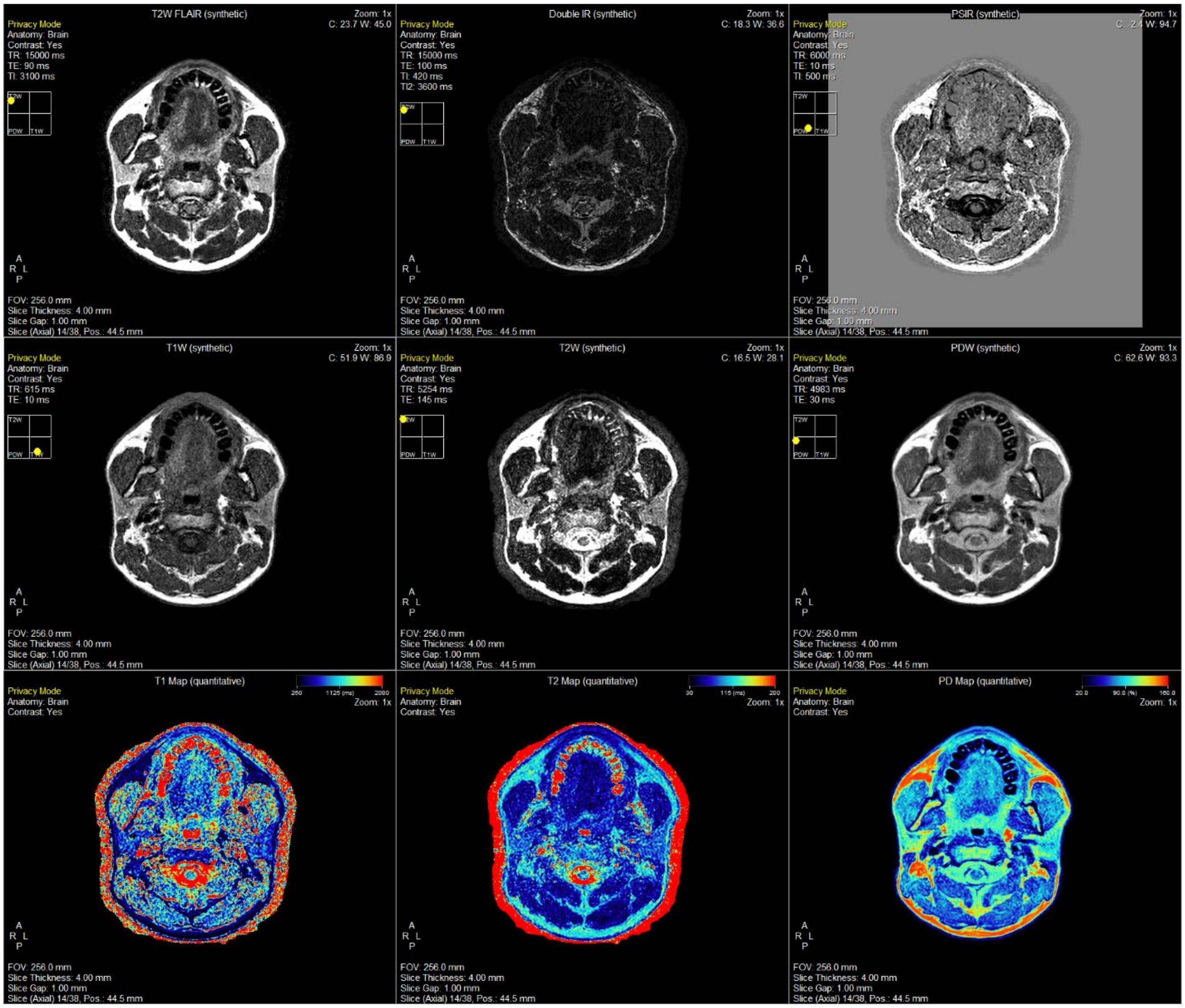

Figure 3-S4. Demonstration of the SyMRI post-processing package offered by SyntheticMR in Volunteer 1 using the fine sequence on the MR-Linac scanner.

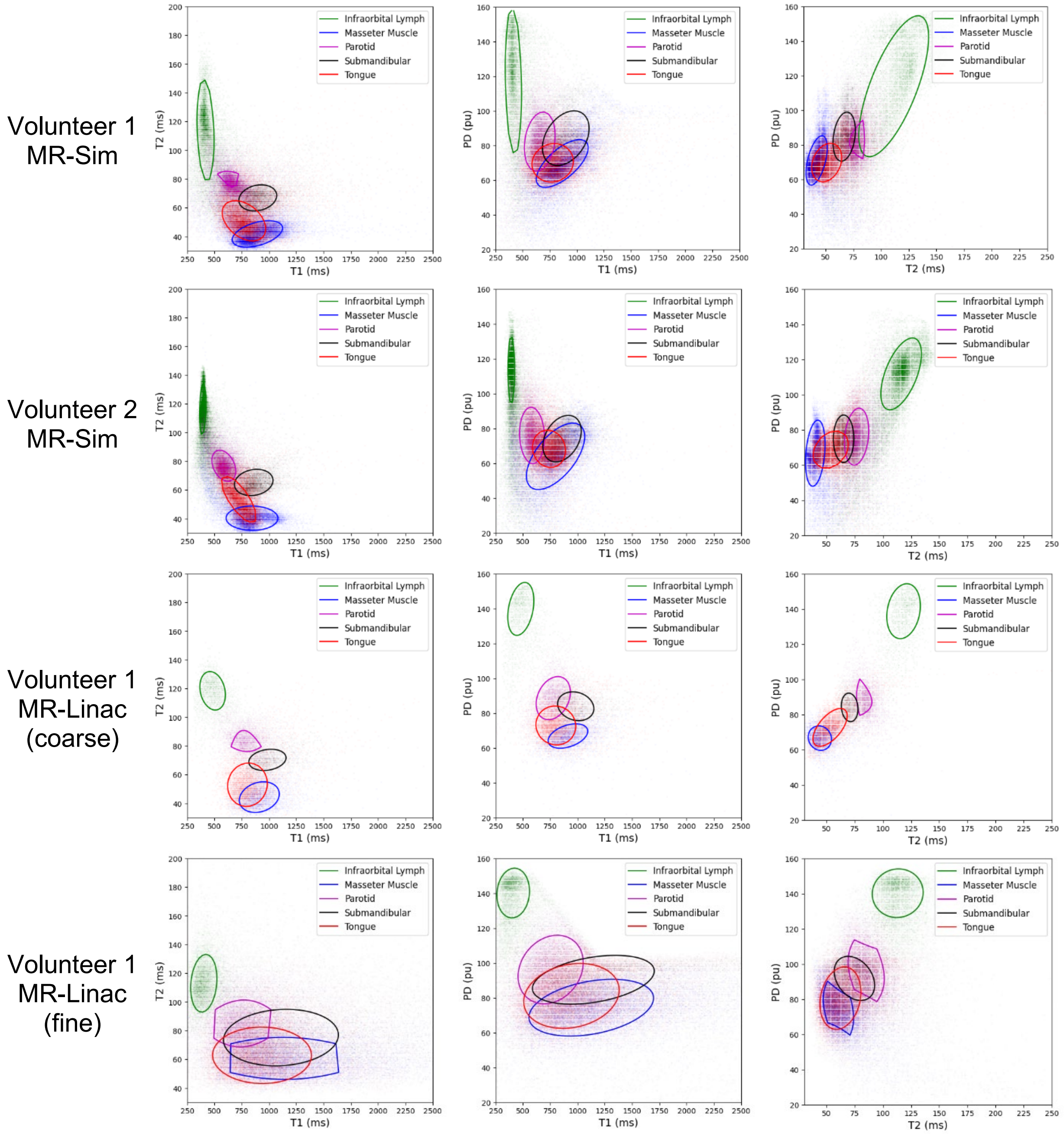


Figure 4-S1. Cluster analysis of normal tissue values for each combination of quantitative parameters (i.e., T1, T2, and PD) on both the MR-Sim and MR-Linac.
